# Survivorship Navigator: Personalized Survivorship Care Plan Generation using Large Language Models

**DOI:** 10.1101/2025.03.27.25324782

**Authors:** Jathurshan Pradeepkumar, Shivam Pankaj Kumar, Courtney Bryce Reamer, Marie Dreyer, Jyoti Patel, David Liebovitz, Jimeng Sun

**Affiliations:** University of Illinois Urbana-Champaign, Urbana, IL; Northwestern University, Chicago, IL

## Abstract

Cancer survivorship care plans (SCPs) are critical tools for guiding long-term follow-up care of cancer survivors. Yet, their widespread adoption remains hindered by the significant clinician burden and the time- and labor-intensive process of SCP creation. Current practices require clinicians to extract and synthesize treatment summaries from complex patient data, apply relevant survivorship guidelines, and generate a care plan with personalized recommendations, making SCP generation time-consuming. In this study, we systematically explore the potential of large language models (LLMs) for automating SCP generation and introduce Survivorship Navigator, a framework designed to streamline SCP creation and enhance integration with clinical systems. We evaluate our approach through automated assessments and a human expert study, demonstrating that Survivorship Navigator outperforms baseline methods, producing SCPs that are more accurate, guideline-compliant, and actionable.

## 1. Introduction

Cancer survivorship spans from diagnosis through the remainder of life, encompassing diverse populations—from individuals managing active cancer to those in remission. According to the Institute of Medicine, cancer survivorship refers to the period following the completion of active treatment, continuing until the end of life[1]. The growing population of long-term cancer survivors, with a current 5-year relative survival rate of 66% in the U.S.[2], faces challenges in transitioning from oncology to primary care, mainly due to the gaps in communication and care coordination[1]. Primary care providers (PCPs), who manage most survivorship care, often lack specialized training in post-treatment complications[3], leading to fragmented follow-up and inadequate patient education about self-care practices[4]. This often leads to redundant or missing essential services[5], negatively impacting health outcomes[6], and imposing substantial financial burdens on both survivors and the healthcare system[7].

Survivorship care plans (SCPs) are standardized tools introduced to bridge this gap and they offer structured, comprehensive guidelines to monitor and improve the health of cancer survivors while facilitating coordinated follow-up care among healthcare providers during the post-treatment or survivorship phase [8]. Recognizing the importance of SCPs, major cancer organizations such as the American Society of Clinical Oncology (ASCO), the National Comprehensive Cancer Network (NCCN), and the American Cancer Society (ACS) have published extensive guidelines for cancer survivorship to enhance their quality [9, 10, 11, 12, 13]. However, their adoption remains critically low in many medical institutions due to several limitations:

- **SCP creation is both time and labor-intensive**: Current SCP creation practices require clinicians to manually extract and synthesize complex patient data to develop treatment summaries, followed by generating personalized recommendations based on existing guidelines. According to [14], clinicians spend an average of over two hours per patient to create a SCP. Such a significant workload makes SCP creation burdensome for busy oncology teams.
- **Adherence to rapidly evolving guidelines**: Cancer survivorship guidelines are frequently updated to reflect emerging research and therapeutic advances. This introduces challenges in maintaining real-time alignment with the latest standards.
- **Interoperability gaps hinder integration**: Most SCPs exist as standalone documents that are not well-integrated into electronic health record (EHR) systems.

**Figure 1:**
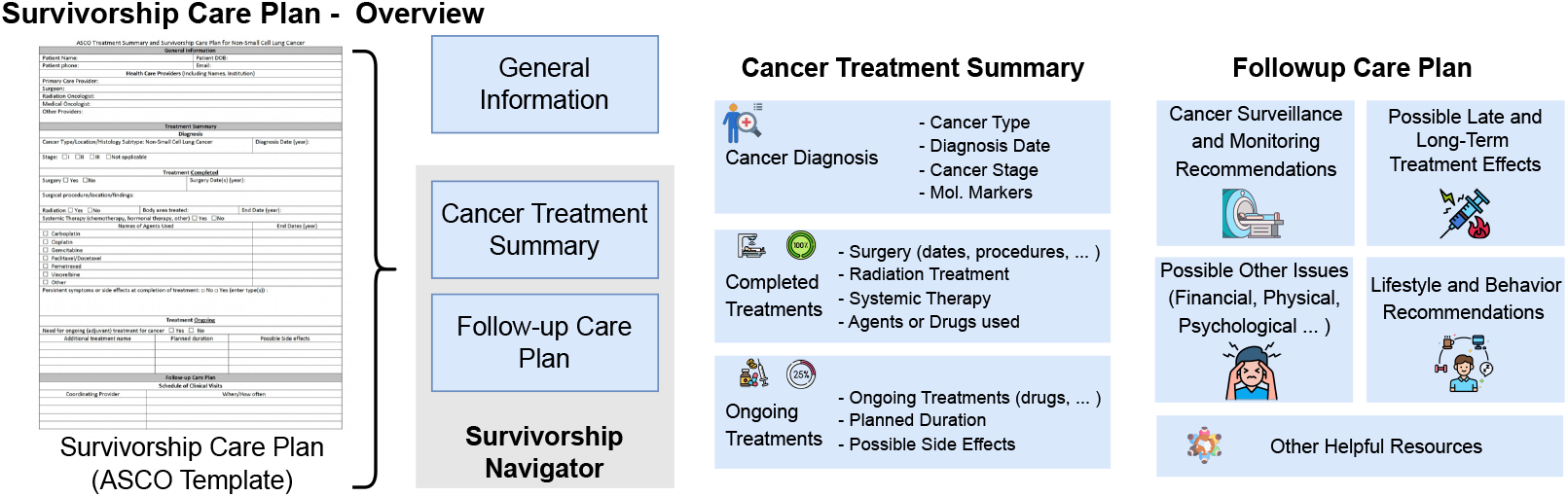
Overview of an SCP based on the template proposed by ASCO[10]. An SCP consists of three sections, where detailed components of the Cancer Treatment Summary and Follow-Up Care Plan are shown on the right.

Large language models (LLMs) have demonstrated remarkable capabilities in various medical tasks, including clinical note generation[15], decision support[16], medical question answering[17], radiology report generation[18] and patient communication[19]. However, the application of LLMs to address challenges in cancer survivorship care, particularly the generation of SCPs remains unexplored. In this paper, we conduct the first systematic investigation of LLMs for survivorship care, with a focus on overcoming the inefficiencies of manual SCP creation. Our proposed *Survivorship Navigator* overcomes the time- and labor-intensive nature of manual SCP creation by automating the synthesis of patient-specific treatment summaries (e.g., chemotherapy regimens, radiation details) and guideline-aligned follow-up care recommendations. Retrieval augmented generation (RAG)[20] is adopted to ground the follow-up care recommendations on existing guidelines to reduce hallucinations. We introduce an automated knowledge base creation pipeline to ensure our framework remains aligned with rapidly evolving guidelines with minimal manual effort. Finally, to enable seamless integration into EHR systems such as EPIC[21], we combine our SCP generator with a Fast Healthcare Interoperability Resources (FHIR)-based interface, bridging the gap between standalone SCPs and actionable clinical workflows. We validate our framework through a multi-stage evaluation consisting of automated and human evaluations.

## 2 Preliminary

### 2.1 Survivorship Care Plan

Several leading cancer organizations have developed templates for SCPs, and in this study, we adopt the SCP template proposed by the ASCO[10]. As illustrated in Figure 1, an SCP is structured into three main sections: (1) General information, (2) Cancer treatment summary, and (3) Follow-up care plan. The general information section typically includes essential demographic details about the patient and information about their healthcare providers, including provider roles and contact details. The cancer treatment summary section presents a comprehensive overview of the patient’s cancer history, covering details such as cancer diagnosis (e.g., cancer type, stage, diagnosis date, molecular markers), completed treatments (e.g., surgeries, radiation therapy, systemic therapies, administered drugs or agents), and ongoing treatments (e.g., continuing medications, planned treatment durations, and potential side effects). The follow-up care plan offers guideline-based recommendations for survivorship, organized into five key tasks (see Figure 1): (1) Cancer surveillance and monitoring (*τ*_1_), (2) Possible late and long-term treatment effects (*τ*_2_), (3) Possible other issues (e.g., financial, psychological)(*τ*_3_), (4) Lifestyle and behavioral recommendations (*τ*_4_) and (5) Helpful resources (*τ*_5_) such as educational materials and patient support groups. Although these templates can be comprehensive, their manual completion by clinicians at discharge is time-intensive and prone to inaccuracy, leading to inconsistent implementation. For this study, our proposed Survivorship Navigator focuses on automating the cancer treatment summary and follow-up care plan sections using de-identified patient data 𝒳.

**Figure 2:**
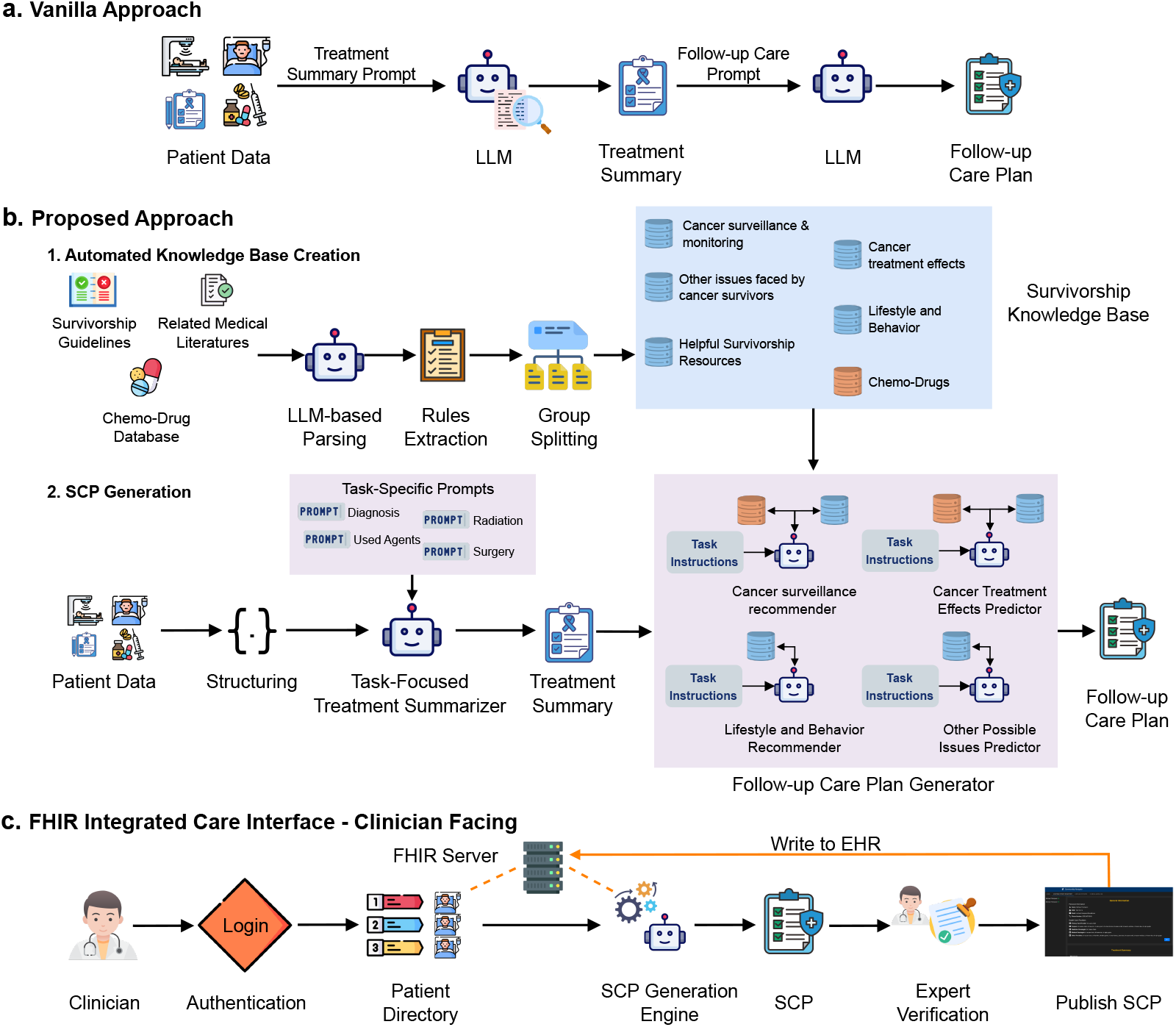
Overview of Survivorship Navigator. (a) Vanilla approach, utilizing a two-stage LLM-based prompting method for SCP generation. (b) Proposed Survivorship Navigator framework, where (b.1) depicts the automated survivorship knowledge base construction, and (b.2) illustrates SCP generation using patient data and the knowledge base. (c) FHIR-integrated care interface, outlining the clinician-facing user workflow.

## 3 Methodology

A straightforward approach to utilizing an LLM for SCP generation is to directly prompt it along with a template to produce the treatment summary and follow-up care plan (see Figure 2a). However, this method presents several challenges: (1) LLMs often omit specific treatment details, (2) they lack specialized domain knowledge, and (3) the generated follow-up care plans tend to be generic rather than patient-specific. In our framework, we propose enhancements to optimize LLM adaptation for SCP generation to address these limitations. The proposed Survivorship Navigator consists of three key modules: (1) Automated survivorship knowledge base creation, (2) SCP generation, and (3) FHIR integrated care interface.

### 3.1 Automated Survivorship Knowledge Base Creation

We adopt a RAG-based approach to ground the recommendations in SCP on existing guidelines and mitigate hallucinations. The first step in our framework is constructing a survivorship knowledge base (see Figure 2b) that extracts and vectorizes information from published survivorship guidelines, relevant medical literature, and drug databases. As new technologies and therapeutics emerge, cancer survivorship guidelines are frequently updated to reflect the latest clinical recommendations. Moreover, individual institutions often utilize distinct guidelines or possess their own documented protocols. Consequently, maintaining a static knowledge base is impractical, and creating such resources typically requires manual effort to develop document-specific parsing and preprocessing algorithms.

Recent works have demonstrated the efficacy of LLMs in processing multimodal data [22] and extracting structured information from scientific texts [23]. We leverage these capabilities to automate the construction of our knowledge base (see Figure 2b). For document *d∈* 𝒟_guidelines_ we parse text and images using multimodal LLMs (e.g., GPT-4o [22]) and extract rules ℛ_*d*_ by prompting. The extracted rules are categorized by task *τ*_*i*_, embedded via OpenAI’s text-embedding-3-small model and stored in task-specific vector datastore 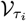 . This process results in the creation of five vector datastores, each corresponding to a specific task within the follow-up care plan(see Figure 2b). We employ semantic similarity-based retrieval followed by reranking using ColBERTv2[24].

General-purpose LLMs often lack domain-specific knowledge, particularly regarding medications, their clinical applications, and associated side effects. To enhance the personalization of follow-up care plans, it is essential to incorporate detailed information about the medications used by cancer survivors, especially for tasks such as cancer surveillance and monitoring recommendations, and predicting potential late and long-term treatment effects. To address this limitation, we mine a chemotherapy database^1^ to construct a chemo-drug datastore 𝒟_drugs_ with following drug information {description, side-effects, drug effect surveillance}. During follow-up care plan recommendations, we first extract drug entities from patient data and then employ lexical matching to retrieval.

### 3.2 Personalized Survivorship Care Plan Generation

As shown in Figure 2b.2, our survivorship care plan generation pipeline comprises two modules: (1) Task-focused treatment summarizer and (2) Follow-up care plan generator.

#### Task-focused treatment summarizer

The ASCO SCP template provides a structured format for cancer treatment summaries [10], requiring the extraction of fine-grained treatment details. For example, in the case of surgical procedures, the model must accurately identify the type of surgery performed along with the date, the anatomical site, and relevant clinical findings. In our initial experiments, we directly prompted an LLM to generate treatment summaries. However, we observed that the model frequently failed to extract certain treatment details, likely due to limited domain knowledge and inconsistent entity recognition (Results presented in Section 4.2). To address these challenges, we employ a two-step process. First, we preprocess and structure the patient data𝒳 , by prompting LLM to convert unstructured data into a structured JSON format: 𝒳_struct_ = LLM( , 𝒳*ϕ*_struct_), where *ϕ*_struct_ denotes structuring prompt. Next, we generate the treatment summary using a task-specific prompting approach formulated as: *S*_*i*_ = LLM(𝒳_struct_, *ϕ*_*i*_, *ϵ*_*i*_), where *ϕ*_*i*_ represents task-specific instructions and *ϵ*_*i*_ consists of examples to enable incontext learning[25]. Here, each task corresponds to the extraction of specific treatment-related information, such as diagnosis, surgical procedures, radiation therapy, ongoing treatments, etc. The effectiveness of this approach is demonstrated in the results presented in Section 4.2.

#### Follow-up care plan generator

As illustrated in Figure 2b, we employ task-specific (*τ*_*i*_) workflows combined with RAG to generate a personalized follow-up care plan. Certain tasks, including cancer surveillance recommendations and prediction of late and long-term treatment effects, require retrieving relevant information from both the chemodrug datastore and the corresponding task-specific datastore. For example, generating appropriate cancer surveillance recommendations requires detailed knowledge of the medications used during treatment. Certain chemotherapy agents or radiation therapies may necessitate specific follow-up tests or regular monitoring due to their potential late effects. The generation process consists of three key steps:

#### Drug information retrieval

Given the treatment summary, we first extract all administered drugs and medications and retrieve their relevant details from the chemo-drug datastore (*R*_drug_).

#### Task-Specific Knowledge Retrieval

The retrieved drug information is then combined with the treatment summary to query the appropriate task-specific datastore for retrieval: 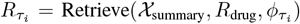, where𝒳 _summary_ represents the treatment summary, and 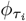 denotes the prompt for the specific follow-up care task.

#### Follow-up care recommendation

The aggregated information is used to prompt the LLM, generating structured, evidence-based follow-up care recommendations. 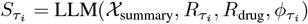

For lifestyle recommendations and prediction of other issues, retrieval is performed only from the task-specific datastore, as these tasks do not require chemo-drug information.

### 3.3 FHIR Integrated Care Interface

To facilitate seamless integration with existing EHR systems, we propose a FHIR-integrated care interface, as illustrated in Figure 2c, which outlines the workflow of the clinician-facing interface. Clinicians first authenticate to access patient records within their clinic. The system then retrieves patient data from the hospital’s EHR system via an FHIR server, which is processed by the SCP generation engine to construct the SCP. The system functions as a clinical assistant, providing verification functionalities that allow clinicians to review and validate the generated SCP for accuracy before finalization. Once verified, the SCP is published in the user interface and written into the EHR system. Patients also will have a dedicated interface to access their SCP upon authentication. Additional features, such as appointment scheduling, follow-up tracking, and SCP updates throughout the survivorship phase, can further enhance functionality and continuity of care.

## 4 Experiments and Results

### 4.1 Experiment Setup

#### Dataset

There are currently no existing benchmarks or ground truth datasets for evaluating LLMs on SCP generation. In a prior small-scale study, we conducted a user study on 10 synthetic patient cases to assess feasibility, presented in [26]. For our experiments, we used the CORAL dataset – a fine-grained, expert-annotated collection of 40 de-identified breast and pancreatic cancer progress notes[27]. While CORAL provides annotations for various clinical entities, such as symptoms and medications, it lacks specific annotations for the structured treatment summary in SCP, which are essential for evaluating our framework. To address this gap, we created a tailored annotation set for SCP generation. The annotation process involved two stages. First, GPT-4o was prompted with each clinical note and its existing annotations to generate an initial set of treatment-related annotations, serving as a reference for human annotators. Subsequently, two domain experts manually reviewed and refined these annotations, producing a high-quality gold standard dataset for evaluating treatment summary extraction.

#### Evaluation Process

We systematically evaluated the capabilities of LLMs in SCP generation and assessed the impact of our proposed framework improvements compared to a baseline vanilla approach using three key evaluation methods: (1) treatment extraction performance using natural language processing (NLP) metrics, (2) LLM-based A/B testing, and (3) human expert evaluation. For this study, we used a combination of open-source and proprietary LLMs, including Mistral 7B, Llama 3.1 8B, Gemma 2 9B, GPT-4, and GPT-4o. Proprietary models were accessed via HIPAA-compliant API calls through Azure. A/B testing and human expert evaluations were conducted using Gemma 2 9B, GPT-4, and GPT-4o.

First, we quantified the ability of LLMs to extract relevant cancer treatment information and generate structured treatment summaries using BLEU-4 with smoothing[28], ROUGE-1[29], and Exact Match (EM) F1 scores. In A/B testing, an LLM (GPT-4o), prompted to act as an oncologist, compared SCPs generated by the vanilla LLM-based approach and our proposed method. The LLM evaluated the SCPs based on comprehensiveness, accuracy, actionability, and personalization, selecting the preferred version.

**Figure 3:**
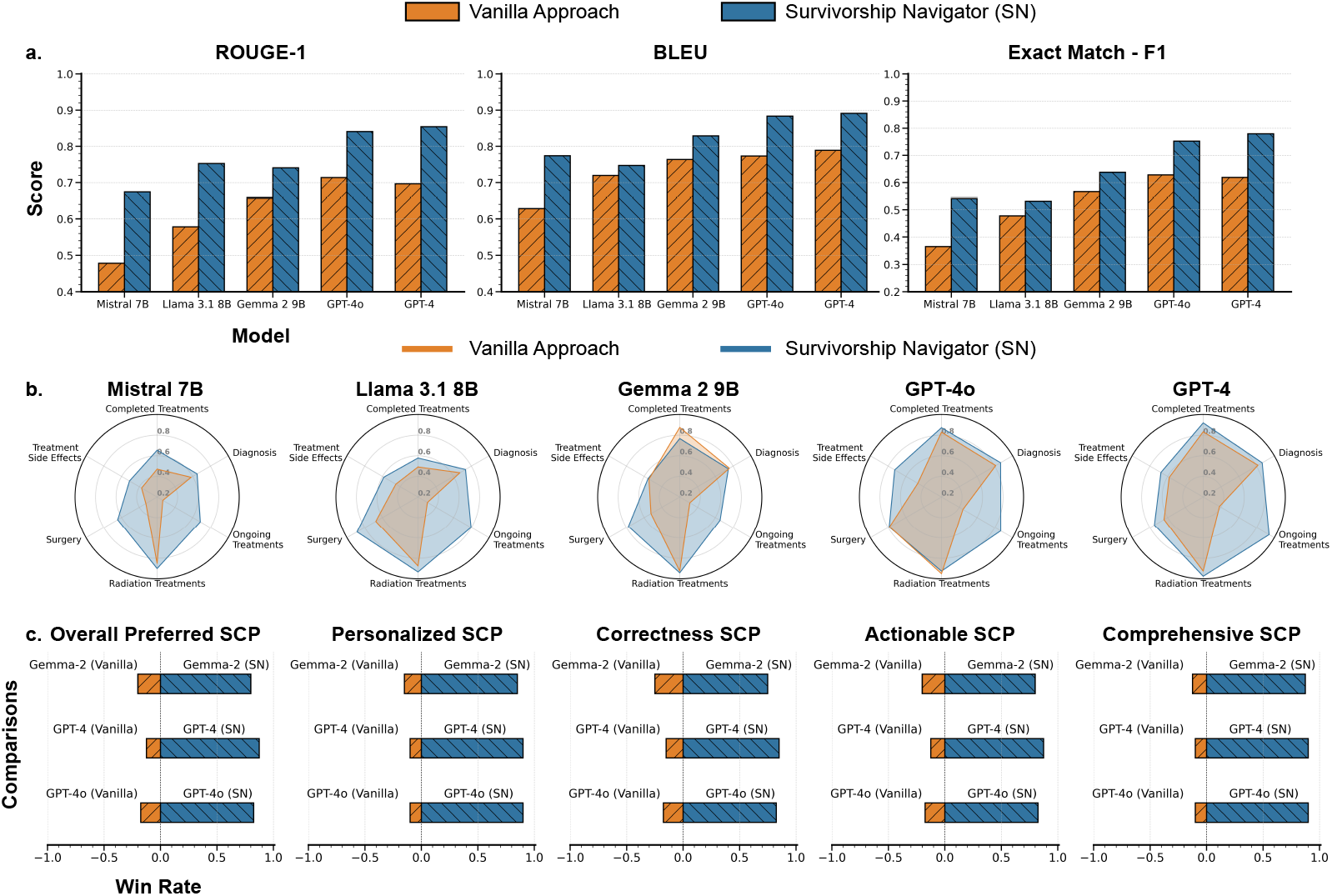
Automated assessment results. (a) Comparison of treatment information extraction performance between the vanilla approach and our method using NLP metrics. (b) Task-wise ROUGE-1 scores across all LLMs, , where tasks correspond to different treatment information types, such as diagnosis, surgeries, and ongoing treatments. (c) LLM-based A/B testing results, evaluating SCP quality across multiple dimensions.

Finally, a domain expert assessed SCPs generated by LLMs. This assessment consisted of two components: (1) treatment information extraction performance, measured by the proportion of correct, incorrect, and missed extractions, and (2) a structured user study where SCPs were rated across multiple dimensions on a 5-point scale. The evaluated dimensions included correctness, faithfulness (alignment with guidelines or retrieved-context), actionability, reasoning accuracy, personalization, clarity and understandability, and comprehensiveness.

### 4.2 Performance Evaluation of Cancer Treatment Extraction

We conducted an automated quantitative evaluation to compare cancer treatment information extracted by different LLMs against our manually annotated references. As outlined in the previous section, we employed three quantitative metrics: BLEU, which emphasizes precision; ROUGE-1, which is more recall-oriented; and EM-F1, which strictly measures lexical matches by computing precision, recall, and their harmonic mean. EM-F1 is particularly strict, penalizing even minor phrasing variations, resulting in lower scores.

The overall performance of treatment information extraction across all LLMs, including a comparison of their vanilla versions and our enhanced variants, is presented in Figure 3a. Task-specific prompts combined with examples consistently improved performance over the vanilla approach across all models and metrics. Notably, significant improvements were observed in smaller open-source models, where their performance, with proposed improvements, matched that of vanilla variants of large proprietary models such as GPT-4 and GPT-4o. A larger increase in the ROUGE-1 score across all LLMs suggests that the vanilla approach frequently misses treatment-related information, whereas our method extracts it more effectively (better recall). GPT-4 and GPT-4o, when integrated with our framework, achieved ROUGE-1 and BLEU scores above 0.8 and an EM-F1 score exceeding 0.7.

**Table 1:** Human evaluation results for cancer treatment information extraction.

| Model | Correct Extractions % | Incorrect Extractions % | Missing Extractions % |
| --- | --- | --- | --- |
| Gemma 2 9B | $0.7111 \pm 0.1581$ | $0.2426 \pm 0.1598$ | $0.0463 \pm 0.0885$ |
| GPT-4 | $0.8640 \pm 0.1182$ | $0.1172 \pm 0.1048$ | $0.0188 \pm 0.0490$ |
| GPT-4o | $0.9015 \pm 0.1244$ | $0.0900 \pm 0.1171$ | $0.0085 \pm 0.0292$ |

To assess the capability of LLMs in extracting different types of treatment information, we analyzed their task-specific performance, as shown in Figure 3b. Our Survivorship Navigator consistently outperformed the vanilla approach across all tasks, with significant improvements in extracting ongoing treatments, treatment side effects, and surgical interventions. Overall, the quantitative results indicate that our framework enhances cancer treatment extraction for SCP generation and consistently improves LLM performance.

However, NLP-based extraction methods have inherent limitations. To address these, we conducted a human evaluation of the treatment extractions. A domain expert manually assessed each extraction generated by our approach using Gemma-2 9B, GPT-4, and GPT-4o, categorizing them as correct, incorrect, or missing. The mean percentage of extractions in each category across all patients is presented in Table 1. The results indicate that GPT-4 and GPT-4o achieved over 85% accuracy in correct extractions, with all three models exhibiting low rates of missing extractions. However, the smaller Gemma-2 9B model achieved only 71.1% accuracy, highlighting its limitations in treatment extraction. These findings provide a more reliable assessment of LLM performance in extracting treatment information.

Despite the overall effectiveness of our approach, several limitations were identified during the analysis. First, the LLMs occasionally hallucinate dates or misattribute events when specific temporal information is missing from the clinical notes. For example, in Subject 26, where no diagnosis date was provided, the model incorrectly inferred the diagnosis date as the chemotherapy start date. Second, the current framework does not handle complex cases involving multiple cancer diagnoses. It primarily focuses on the primary cancer under discussion, often overlooking additional malignancies. In the case of Subject 26, who had colon, endometrial, and breast cancer, the model extracted information only for breast cancer, omitting the others. Future work should explore more sophisticated methods capable of handling such complex clinical scenarios to improve the robustness of treatment information extraction.

### 4.3 Evaluation of SCP Generation via LLM-Based A/B Testing

To evaluate the quality of LLM-generated SCPs across key dimensions: personalization, correctness, actionability, and comprehensiveness, we employed an A/B testing strategy with an LLM-based judge. Specifically, we used GPT-4o to compare SCPs generated by the vanilla approach against those produced using our Survivorship Navigator framework. In this evaluation, GPT-4o was prompted to select the superior SCP in each category without any prior knowledge of its origin. Additionally, the LLM judge determined the overall preferred SCP by selecting the most favorable candidate across all criteria and the evaluation results were output in a structured JSON format. Figure 3c presents the results, measured by win-rate across different dimensions. The findings demonstrate that SCPs generated using the Survivorship Navigator were consistently preferred over the vanilla approach, exhibiting better personalization, comprehensiveness, correctness, and actionability.

### 4.4 Human Expert Evaluation

In order to evaluate the quality of LLM-generated SCPs (Gemma 2, GPT-4, and GPT-4o with our Survivorship Navigator) and gain expert insights, we conducted a human user study focusing on the follow-up care plan section using 20 breast cancer progress notes. A clinical fellow assessed the SCPs using a 5-point scale across multiple dimensions based on the following criteria:

- Correctness: Assesses the factual accuracy of the follow-up care recommendations.
- Faithfulness: Evaluates whether the recommendations align with clinical guidelines, verified against retrieved context from the survivorship knowledge base.
- Actionability: Measures the practical applicability of follow-up care recommendations, specifically for cancer surveillance and monitoring, as well as lifestyle and behavior recommendations.
- Reasoning accuracy: Models were asked to generate explanations, which were then manually reviewed to assess the quality of justifications provided for each recommendation.
- Personalization: Examines the extent to which the SCP is tailored to an individual patient’s needs rather than relying on generic or template-based responses.
- Clarity: Evaluates the readability, coherence, and understandability of the SCP.
- Comprehensiveness: Focuses on the completeness of the SCP, ensuring all relevant aspects are covered.

**Figure 4:**
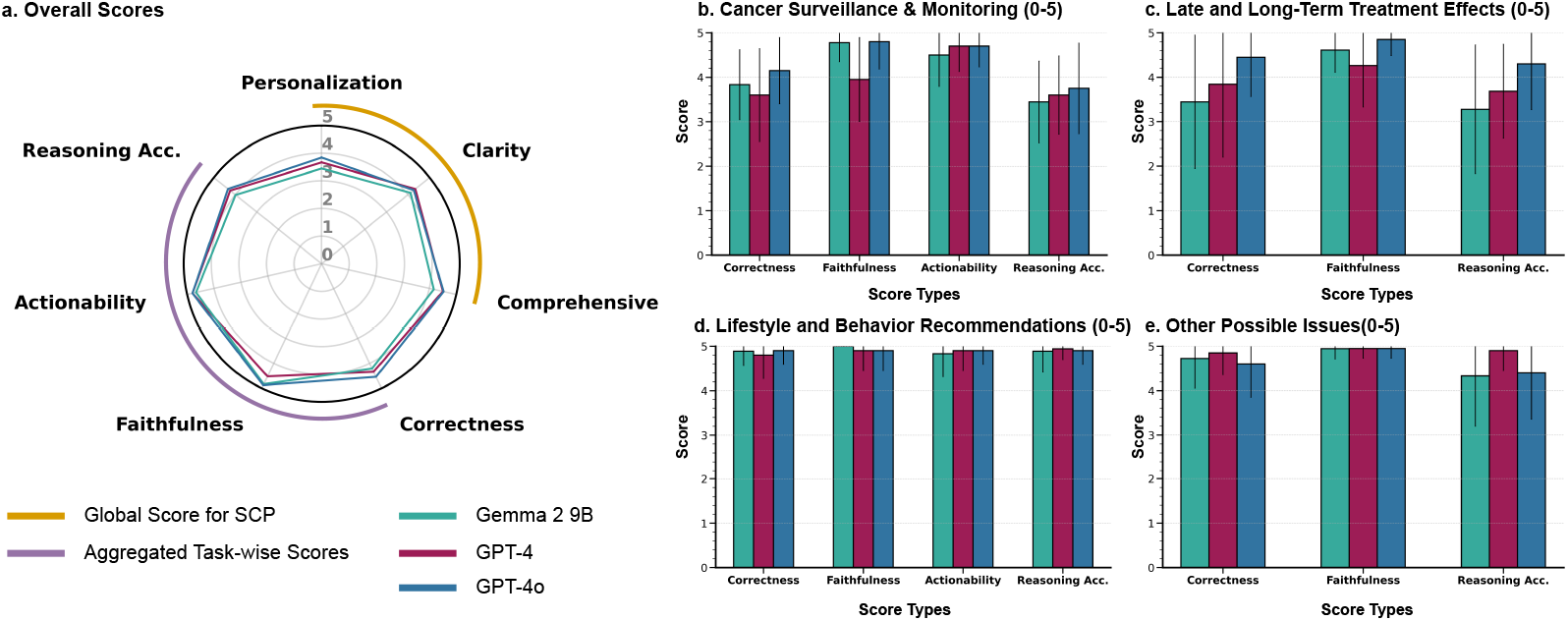
Human expert evaluation of SCPs generated by Survivorship Navigator. (a) Overall scores on a 5-point scale across all three LLMs. (b–e) Task-specific follow-up care evaluation, including (b) cancer surveillance and monitoring recommendations, (c) possible late and long-term treatment effects, (d) lifestyle and behavior recommendations, and (e) other potential survivorship challenges.

For personalization, clarity, and comprehensiveness, the clinical expert assigned an overall SCP score, while task-wise (*τ*_*i*_) scores were obtained for correctness, faithfulness, and reasoning accuracy, which were then aggregated to derive overall performance metrics. The overall results, presented in Figure 4a, indicate that LLM-generated SCPs performed particularly well in actionability, faithfulness, and correctness, highlighting their ability to generate clinically relevant, guideline-aligned, and actionable recommendations. However, personalization received a comparatively lower score (*<* 4), suggesting that further refinements are needed to enhance the individualization of SCPs. A key limitation contributing to this result is the framework’s current inability to address multiple cancer occurrences. For instance, in Subject 26, follow-up care recommendations focused solely on the primary cancer (breast cancer), omitting necessary follow-up for coexisting cancers (colon and endometrial). One barrier to building more robust models is the limited availability of patient data that adequately represents multi-cancer scenarios. Addressing this data gap will be important for future development. Figures 4b, c, d, and e illustrate follow-up care task-specific scores.

Among all models, GPT-4o achieved the highest overall scores across all dimensions, while Gemma-2 9B, a smaller open-source model, demonstrated competitive performance compared to proprietary LLMs. However, Gemma-2 underperformed in personalization and correctness, particularly in cancer surveillance recommendations and predicting late and long-term treatment effects, as shown in Figure 4b and c. Additionally, Gemma-2 demonstrated lower reasoning accuracy compared to GPT models. Despite these limitations, open-source models like Gemma-2 can be enhanced by utilizing domain-specific LLMs (medical LLMs [30]), advanced prompting techniques such as Chain-of-Thought prompting [31], and further fine-tuning using instruction-tuning [32]. These enhancements can be a future direction for improving local LLMs in SCP generation while also ensuring data privacy by enabling deployment within a clinic’s local environment.

The framework demonstrates strong performance in lifestyle and behavior recommendations as well as in identifying other potential issues cancer survivors may encounter during their survivorship, as shown in Figure 4d and e.

However, cancer surveillance recommendations and predicting late and long-term treatment effects appear to be more challenging tasks in follow-up care (see Figure 4b and c). These tasks require higher domain knowledge to generate precise and clinically appropriate recommendations. An important observation is that while both tasks achieve high faithfulness scores, they exhibit lower correctness, suggesting that the model relies heavily on the retrieved context from the knowledge base. Enhancing the knowledge base and improving retrieval accuracy from clinical guidelines could significantly improve correctness and reasoning accuracy, leading to more reliable recommendations.

Considering the task-wise evaluation, the frameworks perform well on lifestyle and behavior recommendations and predicting other possible issues that cancer survivor may face during their survivorship, as shown in Figure 4d and e. Recommending tests and medical checkups for cancer surveillance and predicting late and long-term treatment effects look to be harder tasks in follow-up care (see Figure 4b and c, as these tasks require higher domain knowledge to make better recommendations. One observation is that both tasks achieve higher faithfulness scores but lower correctness and reasoning accuracy, which suggests that these can be improved by improving the knowledge base and enabling better accurate retrievals from guidelines.

## 5 Conclusion

We investigate the capabilities of LLMs in SCP generation and introduce Survivorship Navigator, a framework leveraging LLMs to automate SCP generation, integrating task-specific prompting, RAG, and an FHIR-based interface for seamless EHR integration. Our experiments demonstrate that Survivorship Navigator outperforms the vanilla approach in SCP generation, while a human user study confirms that the generated SCPs are accurate, faithful, and actionable. Future work will focus on enhancing retrieval mechanisms, leveraging domain-specific LLMs, and improving personalization by addressing complex cancer scenarios, ensuring more accurate, scalable, and clinically reliable SCP generation to improve survivorship care.

## Data Availability

All data utilized in this study are available at https://physionet.org/content/curated-oncology-reports/1.0/

## Footnotes

1 https://my.clevelandclinic.org/health/treatments/24323-chemotherapy-drugs

